# Deep learning-based recognition model for surgical phases of minimally invasive hysterectomy: A multicentre retrospective study

**DOI:** 10.64898/2026.05.13.26353100

**Authors:** Ryo Koike, Shin Takenaka, Yukio Suzuki, Hiroki Matsuzaki, Yuichi Harada, Makoto Nakabayashi, Yusuke Hirose, Kenro Chikazawa, Kanae Shimada, Eri Yoshiizumi, Hiroaki Komatsu, Hiroshi Tanabe, Koji Matsumoto

## Abstract

**Objective:** To develop and validate a robust deep-learning model capable of fine-grained phase recognition in total hysterectomy, particularly the complex periuterine dissection phase.

**Design:** Multicentre retrospective observational study.

**Setting:** Japan.

**Sample:** Surgical videos (n = 764) from 43 institutions.

**Methods:** We developed a robust and generalisable deep-learning model for surgical phase recognition in total hysterectomy, applicable to laparoscopic and robot-assisted procedures. Overall, 1,591,334 still images were annotated across nine surgical phases. A convolutional neural network (Xception architecture) was trained on 200 cases using four-fold cross-validation, with institutional separation between training and testing sets.

**Main outcome measures:** Model performance was assessed using accuracy, precision, recall, and F1 score. Subgroup analysis and logistic regression evaluated the association between background clinical factors and recognition accuracy.

**Results:** The model achieved an overall phase recognition accuracy of 0.78 (95% CI: 0.74–0.80), with a precision of 0.75 (95% CI: 0.72–0.78) and a recall of 0.76 (95% CI: 0.74–0.78). Performance was consistent across laparoscopic and robot-assisted procedures and across most surgical phases. Accuracy plateaued after training on 120 cases. No clinical factors significantly impacted performance. Trends toward lower accuracy were observed for cases with cervical myoma and pouch of Douglas adhesions.

**Conclusions:** This model demonstrated high accuracy across diverse institutions and patient backgrounds. Its potential applications include surgical education, real-time intraoperative support, and training efficiency enhancement.

**Funding:** Jmees Incorporated [C2020-127], Japan Agency for Medical Research and Development (AMED) [JP22he0122024]; Japan Society for the Promotion of Science (JSPS) KAKENHI [JP21K12744].

## INTRODUCTION

Hysterectomy—the surgical removal of the uterus—is the most commonly performed gynaecologic procedure worldwide,^1^ with an incidence of approximately 5 per 1,000 women in the United States.^2^ Minimally invasive approaches, including laparoscopic and robot-assisted techniques, now account for over two-thirds of all hysterectomies in the United States,^3^ and over 50% in many other countries.^4^ Given this widespread adoption, there is a growing interest in leveraging artificial intelligence technologies, such as deep learning, to enhance intraoperative understanding and support surgical education in minimally invasive hysterectomy.

Deep-learning models capable of recognising surgical phases are being developed for various surgical procedures.^5–8^ In general surgery, deep learning-based models for surgical phase recognition have shown promise in supporting education and skill assessment.^9–13^ By comparing operative phase durations to expert benchmarks, these models can help identify technical deficiencies and guide targeted training. However, surgical techniques vary widely across countries, institutions, and surgeons. Therefore, developing a robust and generalisable phase recognition model requires training on diverse cases that reflect a wide range of surgical techniques, diseases, and patient characteristics.

Previously reported surgical procedure recognition models for hysterectomy have been limited to small-scale, single-facility, or multi-facility reports that are focused on benign cases and restricted to laparoscopic procedures. Furthermore, in these reports, the dissection around the uterus—the most important step in total hysterectomy—is not divided into detailed procedures but is grouped into one or two steps.^14–17^ In the present study, we built upon these efforts by constructing a large, multi-institutional database encompassing both benign and malignant cases and both laparoscopic and robotic approaches, with a more detailed classification of periuterine dissection phases. The objective of this study was to develop and validate a robust deep-learning model capable of fine-grained phase recognition, particularly for the complex periuterine dissection phase. We hypothesised that (1) a large and diverse dataset would enable detailed phase classification without compromising accuracy; (2) the relationship between dataset size and model performance could be quantitatively assessed to identify the minimum required case volume; (3) patient and disease characteristics would reveal key factors associated with reduced model accuracy; and (4) a unified model could be constructed to function effectively across both laparoscopic and robot-assisted approaches.

## METHODS

This multicentre retrospective study was conducted between October 2020 and March 2025. It employed surgical videos of total laparoscopic hysterectomy (TLH) and robot-assisted laparoscopic hysterectomy (RALH) recorded between October 2017 and May 2022. This study was conducted in accordance with the principles embodied in the 1964 Declaration of Helsinki (as revised in Brazil in 2013). This study protocol was reviewed and approved by the Medical Research Ethics Review Committee of the Japanese Foundation for Cancer Research (registration no.: 2020-375) on 28 December 2020. Written informed consent was obtained from all patients for the use of their surgical videos and clinical data, including commercial use. Patients were not involved in the design, conduct, reporting, or dissemination of this research.

Minimally invasive hysterectomy cases were extracted from a multicentre surgical video database and used to construct a surgical phase recognition model, serving as the training dataset. The inclusion criteria were as follows: (1) minimally invasive hysterectomy for benign or malignant indications, (2) availability of a complete surgical video, and (3) patient consent for data use. The exclusion criteria were incomplete videos, poor quality footage, and missing data; cases meeting these criteria were excluded at the database entry stage. The database included 764 hysterectomy videos from 43 hospitals across Japan. Cases were randomly selected regardless of the surgical approach (laparoscopic or robotic) or pathology (benign or malignant). To ensure representative surgical views, half of the included procedures were purposively selected among those performed by board-certified specialists from the Japan Society of Gynecologic and Obstetric Endoscopy and Minimally Invasive Therapy.

The surgical workflow was divided into nine detailed surgical phases, as follows: (1) setup, (2) retroperitoneal dissection, (3) transection of the utero-ovarian or infundibulopelvic ligaments, (4) bladder dissection, (5) parametrium division, (6) uterine separation, (7) uterine retrieval, (8) vaginal cuff closure, and (9) final check.

A board-certified obstetrician-gynaecologist from the Japan Society of Obstetrics and Gynecology provided lectures on phase definitions (Table 1) to three annotators, including one medical professional.

**Table 1.**
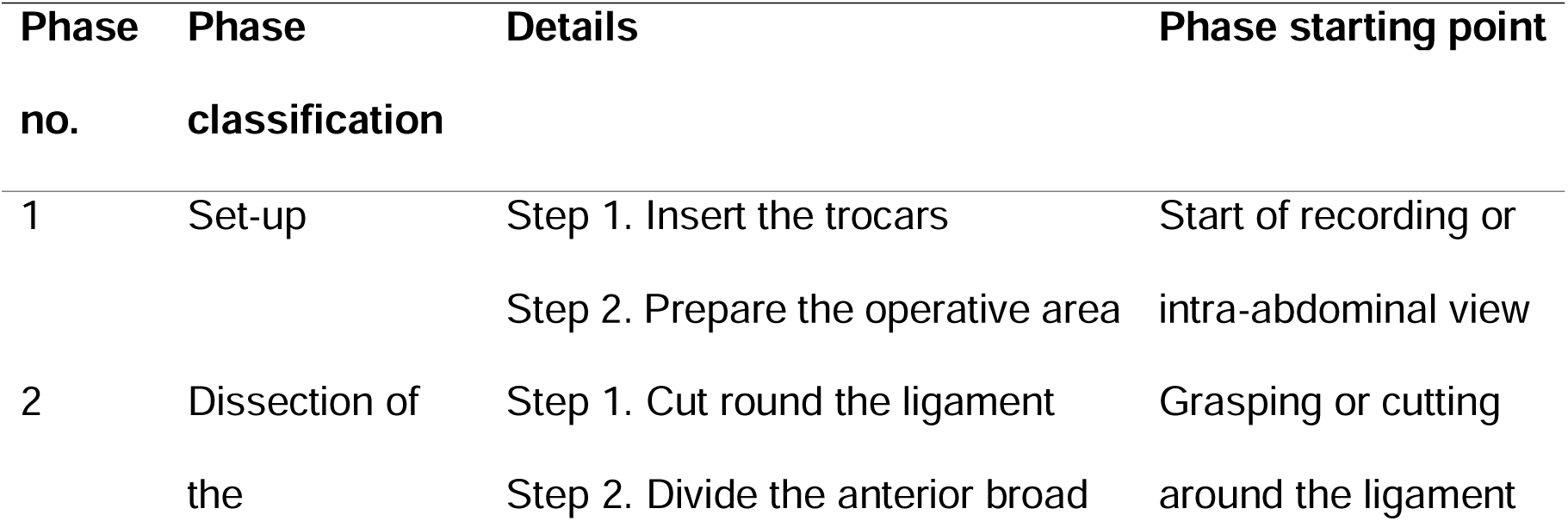
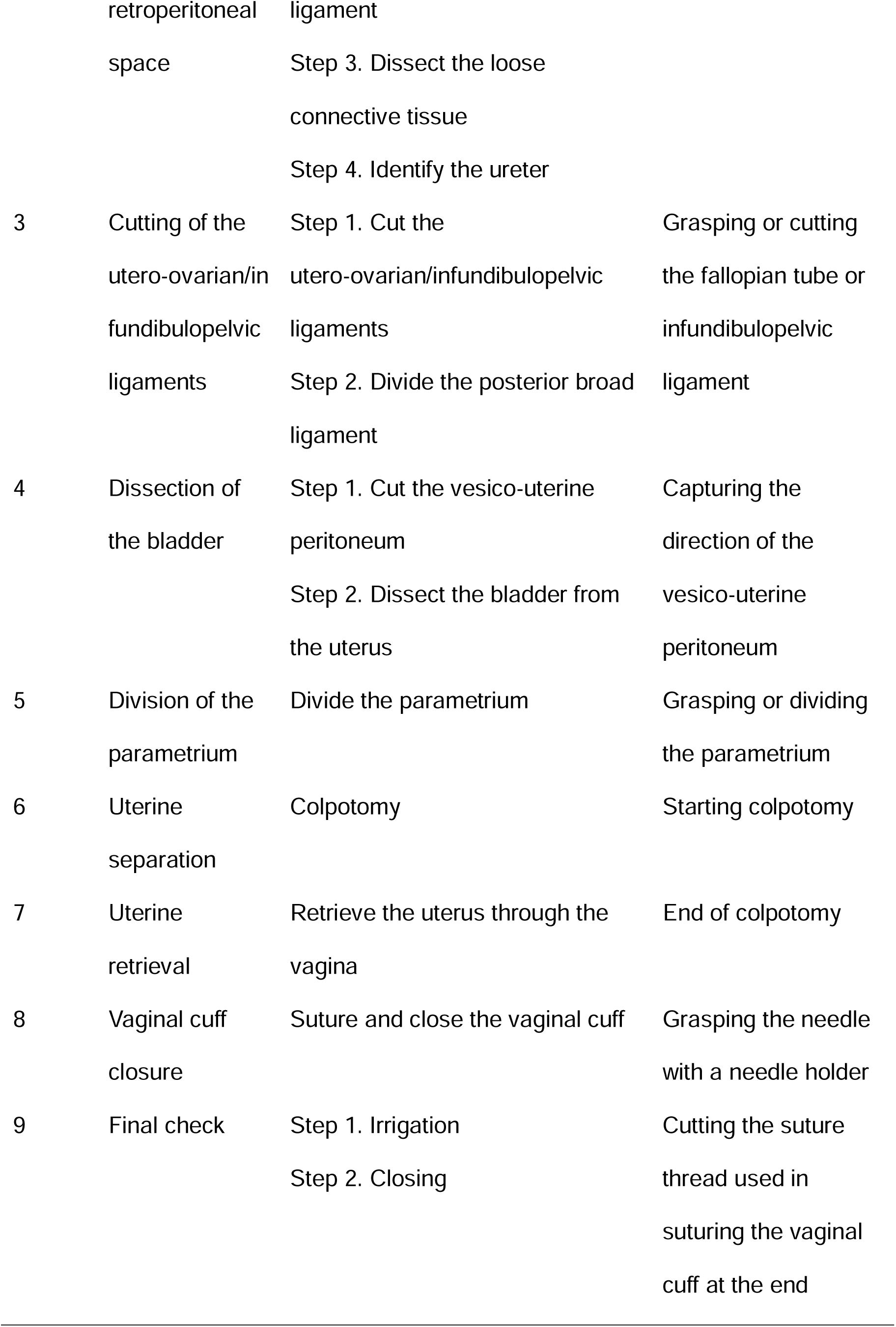
Definitions of the nine surgical phases of total hysterectomy.

All annotators were trained on the annotation software and classification criteria. Still images were extracted from each video at 1-s intervals and labelled with the corresponding surgical phase. Annotations were reviewed and corrected by five Japan Society of Obstetrics and Gynaecology-certified specialists to ensure consistency and accuracy.

A deep-learning model based on the Xception architecture^18^ was trained using the annotated dataset. Xception was selected for its strong performance in complex image recognition tasks. The model parameters are detailed in Table S1.

The model predicted the surgical phase for each still image by selecting the class with the highest probability. Model performance was initially assessed using a fold-out validation approach, where the dataset was progressively expanded to monitor accuracy trends. Once the performance plateaued, the dataset was finalised, and four-fold cross-validation was performed (Figure 1).

**Figure 1.**
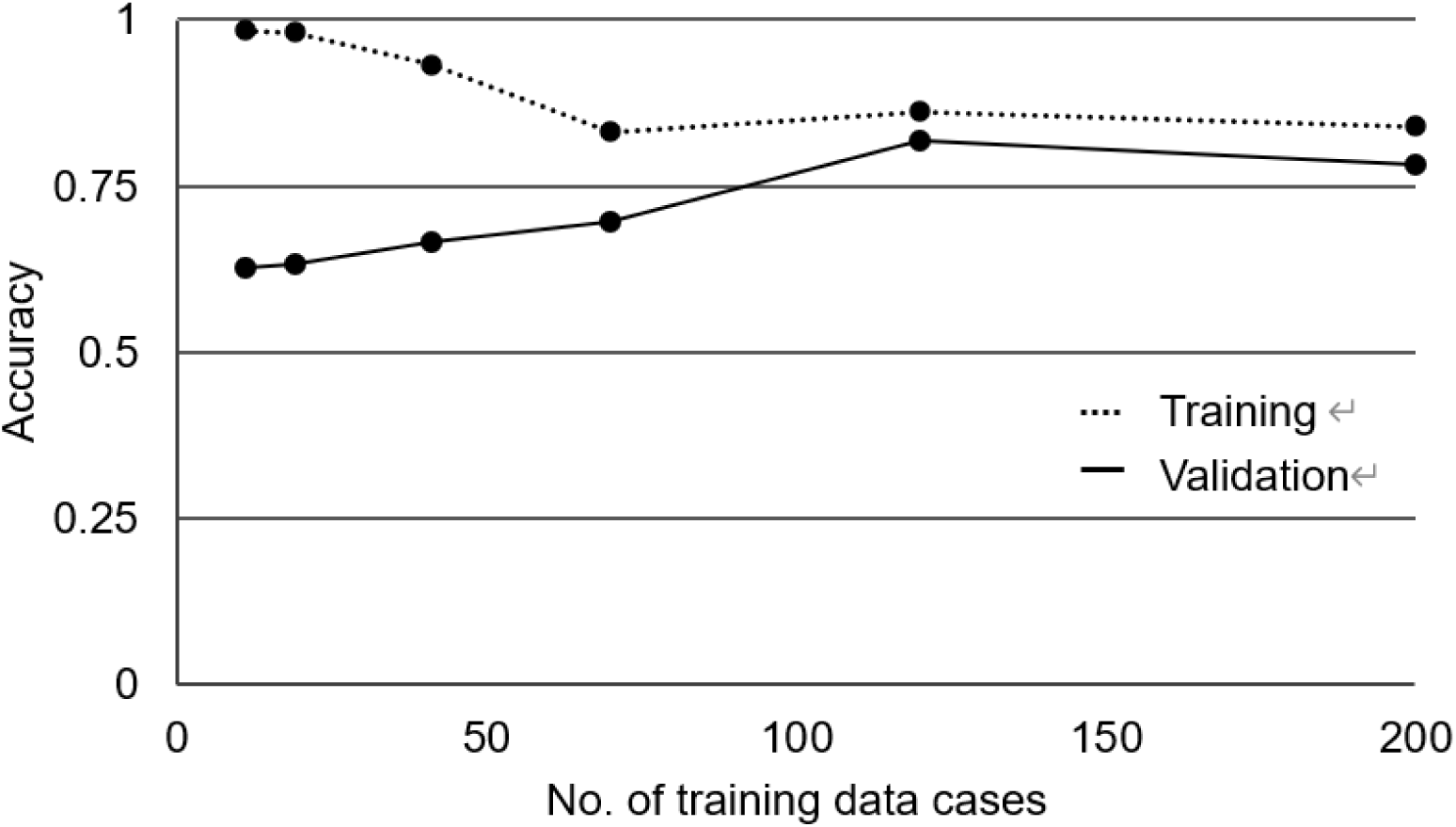
Trends in the accuracy of the deep learning-based surgical phase recognition model for the number of training data cases.

In each fold, three subsets were used for training and one subset was used for testing, ensuring that data from the same facility were not used in both training and testing, to avoid institutional bias.

We could not predict the number of samples required for the model to reach plateau accuracy using a deep-learning approach. Therefore, we gradually increased the number of samples to determine the required number of cases.

The overall accuracy, precision, recall, and F1 score were used to evaluate the machine learning model’s surgical phase classification. These metrics were calculated using the following formulae, where TP refers to true positive; FP, false positive; TN, true negative; and FN, false negative:

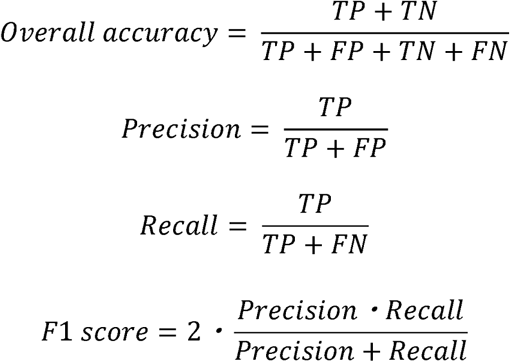

In addition to overall accuracy, phase-specific accuracy was assessed. To determine whether the model could be applied to both TLH and RALH, the accuracy for each surgical approach was calculated separately. We further investigated the clinical factors potentially associated with reduced recognition accuracy and inadequate surgical field exposure. Variables included body mass index (kg/m²), history of abdominal surgery, uterine size (above or below the sacral promontory), presence of cervical myoma ≥3 cm, adhesions in the pouch of Douglas, surgical approach (TLH or RALH), estimated blood loss (mL), surgeon experience (post-graduate year), and post-operative complications defined as Clavien–Dindo grade ≥IIIA or Common Terminology Criteria for Adverse Events v5.0 grade ≥3.

A logistic regression model was used to assess the association between these factors and achieve a phase recognition accuracy of ≥80%. Patient characteristics are presented as mean ± standard deviation and median (interquartile range [IQR]). Statistical analyses were performed using JMP version 17 (SAS Institute, Cary, NC, USA). This study was reported in accordance with the Strengthening the Reporting of Observational Studies in Epidemiology guidelines. This retrospective observational study was not registered in a clinical trial database.

## RESULTS

In total, 1,591,334 still images were annotated across the nine surgical phases as follows: (1) setup, n = 129,412; (2) retroperitoneal dissection, n = 286,825; (3) ligament transection, n = 251,034; (4) bladder dissection, n = 137,680; (5) parametrium division, n = 131,340; (6) uterine separation, n = 85,196; (7) uterine retrieval, n = 163,515; (8) vaginal cuff closure, n = 251,247; and (9) final check, n = 155,085.

To assess the effect of dataset size on model performance, we trained models using incrementally larger subsets of the available cases as follows: 11, 19, 41, 70, 120, and 200 cases. The corresponding accuracy values were 0.62, 0.63, 0.66, 0.69, 0.81, and 0.78, respectively. A steady improvement in accuracy was observed for up to 120 cases, after which performance plateaued and slightly decreased at 200 cases, suggesting diminishing returns from additional data.

The largest dataset, comprising 200 cases, included 1,591,334 annotated frames, covering a wide variety of intraoperative scenarios. These results indicated that the dataset size used in our final experiments was sufficient to achieve stable performance.

Figure 2 shows the confusion matrix for accuracy in each surgical phase.

**Figure 2.**
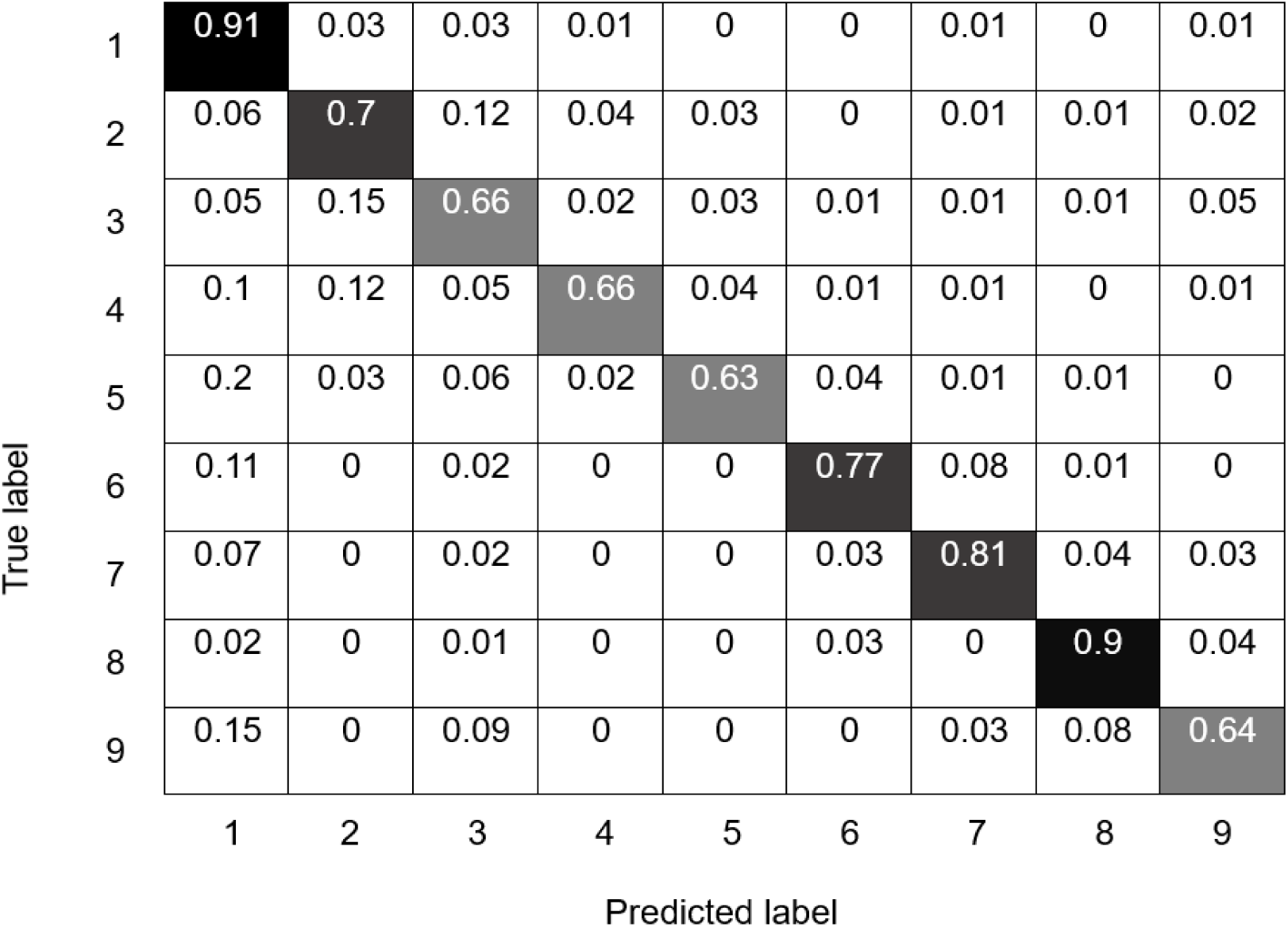
Confusion matrix showing accuracy for each surgical phase

Table 2 shows the model performance for phase classification.

**Table 2.** Precision, recall, F1 score, and accuracy of surgical phase recognition.

| Phase no. | Precision<sup>a</sup> | Recall<sup>b</sup> | F1 score<sup>c</sup> | Accuracy<sup>d</sup> |
| --- | --- | --- | --- | --- |
| 1 | 0.495 | 0.910 | 0.641 | 0.91 |
| 2 | 0.762 | 0.704 | 0.732 | 0.70 |
| 3 | 0.694 | 0.664 | 0.679 | 0.66 |
| 4 | 0.813 | 0.662 | 0.730 | 0.66 |
| 5 | 0.794 | 0.628 | 0.701 | 0.63 |
| 6 | 0.800 | 0.772 | 0.786 | 0.77 |
| 7 | 0.825 | 0.810 | 0.817 | 0.81 |
| 8 | 0.900 | 0.902 | 0.901 | 0.90 |
| 9 | 0.725 | 0.644 | 0.682 | 0.64 |

<sup>a</sup> TP/(TP+FP). <sup>b</sup> TP/(TP+FN). <sup>c</sup> $2 \cdot (\text{Precision} \cdot \text{Recall}) / (\text{Precision} + \text{Recall})$ . <sup>d</sup> (TP+TN)/(TP+FP+TN+FN)
Abbreviations: FN, false negative; FP, false positive; TN, true negative; TP, true
positive

The overall accuracy, precision, and recall were 0.78 (95% CI: 0.74–0.80), 0.75 (95% CI: 0.72–0.78), and 0.76 (95% CI: 0.74–0.78), respectively. Phase-specific accuracies ranged from 0.63 to 0.91. Each phase achieved an accuracy greater than 0.60. The model’s accuracy was 0.78 for TLH cases and 0.78 for RALH cases.

The 200 cases used for training had a median (IQR) age of 47 (43.0–50.0) years. Of these, 173 patients had benign disease and 27 had malignant disease. Disease type included uterine myoma (n = 133), endometriosis (n = 37), cervical intraepithelial neoplasia (n = 15), atypical endometrial hyperplasia (n = 4), endometrial polyps (n = 4), ovarian tumours excluding endometriotic cysts (n = 2), endometrial cancer (n = 25), and cervical cancer (n = 2). Some patients had multiple conditions.

Procedures included 162 TLH and 38 RALH. The median (IQR) specimen weight, operative time, and blood loss were 250.0 (149.7–376.0) g, 164.0 (133.7–214.2) min, and 20.0 (5.0–50.0) mL, respectively. The intraoperative and post-operative complication rates were both 6.0%.

Evaluation of the association between recognition accuracy and clinical factors (Table S2) revealed no significant predictors of reduced deep-learning performance (Figure 3).

**Figure 3.**
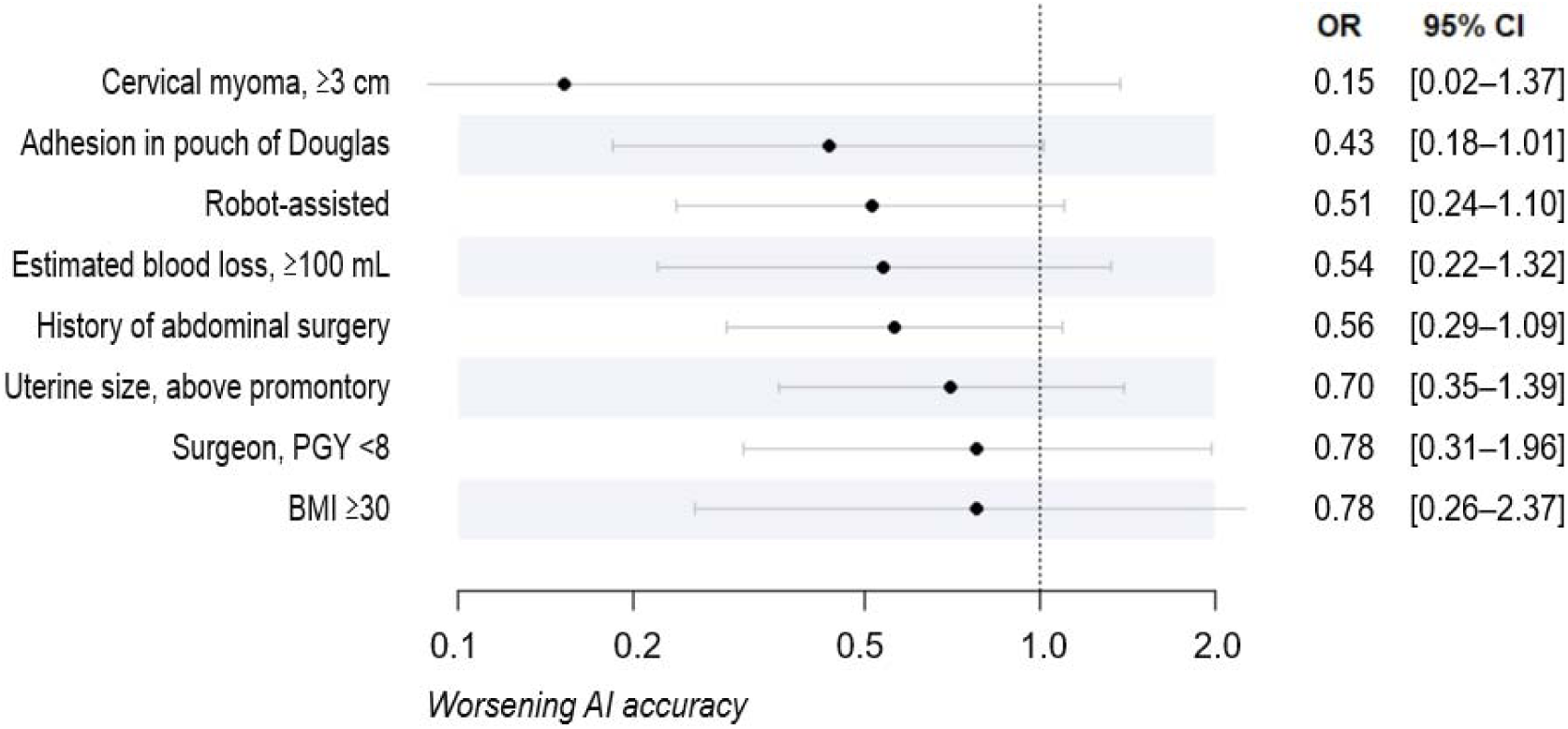
Forest plot of factors associated with worsening deep-learning accuracy. Abbreviations: OR, odds ratio; CI, confidence interval; PGY, post-graduate year; BMI, body mass index; AI, artificial intelligence

Subgroup analysis indicated trends toward lower accuracy in cases with cervical myoma ≥3 cm and pouch of Douglas adhesions.

## DISCUSSION

### Main findings

In this study, we developed a deep learning-based model for surgical phase recognition in minimally invasive hysterectomy, incorporating detailed classification of uterine dissection phases. Using a dataset from a multicentre surgical video database, the model achieved consistently high recognition accuracy across varying institutions, patient backgrounds, surgical skill levels, and device types. The high training and validation accuracy—and the gradual narrowing of the gap between them—suggested strong generalisability across diverse cases.^19^ Additionally, we estimated the minimum number of cases required to stabilise model performance in a multicentre setting based on the performance plateau. No background factors significantly affected model accuracy; however, several clinical trends were observed.

### Strengths and limitations

The strengths of this study include its large and diverse dataset, comprising 764 hysterectomy videos from 43 institutions; these encompass both benign and malignant cases, as well as laparoscopic and robotic approaches. Expert-reviewed annotations and institution-independent validation ensured reliability and generalisability. Another strength is the quantitative assessment of a large dataset relative to model performance, providing a practical benchmark for future research. Finally, we examined the impact of patient and disease backgrounds on recognition accuracy, confirming overall robustness while identifying trends in anatomically complex cases. These features highlight the novelty and clinical relevance of our model.

This study has some limitations. First, all surgical videos were from Japanese institutions, and selection bias may have been introduced owing to non-randomised centre participation. Second, external generalisability remains untested; validation using international data from diverse clinical environments is needed. Third, although the phase definitions used in this study were clinically relevant, alignment with global standards remains uncertain. Finally, the model’s utility in surgical education and skill assessment has not been formally evaluated. Further research is warranted to evaluate its impact in real-world training contexts.

### Interpretation

The overall accuracy of our model was 0.78, although there were differences in accuracy among the nine surgical phases. Dissections in phases 2–5 involve adjacent periuterine structures, making it difficult to delineate phase boundaries. This complexity led to challenges in annotation definition and frequent misclassification among these phases. Moreover, due to the distant view commonly used in phase 1, there were frequent misrecognitions between phase 1 and other phases, particularly phase 9. These factors likely contributed to reduced recognition accuracy. Future improvements may include refining annotation protocols based on intra-abdominal anatomical landmarks and classifying distant-view images as a separate phase. In contrast, phases 1, 6, 7, and 8 consisted of more typical and well-defined scenes, resulting in higher recognition accuracy. This finding is consistent with previous studies reporting high accuracy in phase classification models.^14–17^

Previous studies employing surgical phase recognition models have reported accuracies ranging from 0.76 to 0.93^14–17^; however, these studies were limited by the use of single-centre datasets and relatively small sample sizes. Recently, Levin et al.^17^ developed a model using multi-institutional data, achieving an accuracy of 0.93. However, existing models still present several limitations. Notably, the phase definitions employed in studies by Malpani et al.,^14^ Meeuwsen et al.,^15^ Guédon et al.,^16^ and Levin et al.^17^ consisted of only five to ten broad categories, with many procedural steps grouped under general phases such as ‘setup’ or ‘closure’. In the context of hysterectomy, for example, the periuterine dissection phase—a critical component of the procedure—is frequently not subdivided.

In practice, periuterine dissection involves a series of stepwise manoeuvres to detach the uterus from surrounding anatomical structures. These typically include (1) dissection of the retroperitoneal space, (2) transection of the utero-ovarian or infundibulopelvic ligaments, (3) mobilisation of the bladder, and (4) division of the parametrium. When these steps are aggregated into one or two phases, determining which specific step is the most time-consuming—an essential consideration in surgical evaluation and optimisation—becomes impossible.

In contrast, our model was developed using a large, multicentre dataset comprising hundreds of cases and encompassing both laparoscopic and robotic hysterectomies. This allowed for a finer-grained phase classification, including detailed subphases of uterine dissection. Despite the increased number of defined phases, our model maintained high recognition accuracy, thereby enabling more precise procedural analysis than previously reported systems.

Importantly, cross-validation using institution-independent test sets confirmed the robustness and generalisability of our model. To our knowledge, this is the first surgical phase recognition system that supports both laparoscopic and robot-assisted hysterectomy. Our dataset included a 4:1 ratio of laparoscopic to robotic cases, reflecting the current distribution of surgical practice in Japan. The comparable accuracy observed across both modalities suggests that surgical instrument type had minimal influence on model performance, possibly due to shared visual characteristics between laparoscopic and robotic views. This dual-modality capability may serve as a foundation for expanding phase recognition systems into other surgical domains.

Model performance stabilised beyond 120 training cases, with no further gains observed at 200 cases. This plateau suggests the achievement of model maturity and the elimination of overfitting. Thus, 200 cases were adopted for model construction. Although few studies have addressed required sample sizes for multicentre surgical phase models, Kitaguchi et al.^20^ reported using 300 cases for sigmoidectomy. Our findings offer a practical benchmark; however, annotation consistency and image quality also influence performance. We believe that this report will provide insights into setting the number of samples in similar future studies.

We also examined eight background factors for associations with recognition model accuracy. Although no factors reached statistical significance, trends toward reduced accuracy were observed in cases with cervical myoma and adhesions in the pouch of Douglas. These anatomical features may obscure landmarks or alter dissection planes, complicating phase identification even for human observers. The model demonstrated overall robustness; however, performance in anatomically complex cases may benefit from targeted training on such subgroups. Future studies should focus on developing specialised submodels or context-aware algorithms to maintain high accuracy in complex scenarios.

Beyond recognition performance, the model offers practical utility in surgical education by addressing the limitations of traditional skill assessment methods, which often rely on subjective evaluation, are inconsistent, and require expert human resources. By providing automated, phase-specific feedback without manual annotation, the model enables early identification of technical deficiencies and supports more focused and efficient learning. In addition, the model has the potential to facilitate quantitative comparisons of each phase’s duration and quality between trainees and expert-level benchmarks, should such reference data become available.^21^ This capability may enable targeted practice by highlighting the phases in which trainees underperform, ultimately accelerating skill acquisition. When used continuously, the system could allow trainees to visualise their personal learning trajectories over time, offering objective self-assessment. This function would be particularly beneficial in resource-limited settings where access to expert mentorship may be constrained, contributing to the standardisation and equalisation of surgical education across institutions. Ultimately, this may contribute to improved patient outcomes.

## CONCLUSION

We developed a robust, generalisable deep-learning model for surgical phase recognition in total hysterectomy that demonstrated high accuracy across modalities and institutional settings. The proposed model represents an important step toward automated, standardised surgical analysis. Future improvements may be achieved by incorporating more anatomically complex cases and validating the model’s educational applications.

## Acknowledgements

The authors thank the following individuals for their cooperation in collecting surgical videos and patient information: Moriwaki M, Obihiro Kosei Hospital; Tamate M, Sapporo Medical University Hospital; Ota H, Teine Keijinkai Hospital; Tamura R, Aomori Prefectural Central Hospital; Kasai A, Hachinohe City Hospital; Baba T, Iwate Medical University School of Medicine; Shikama A, University of Tsukuba Hospital; Komatsu H, Tone Chuo Hospital; Kamozawa C, Tochigi Cancer Center; Oishi H, Central Hospital of the National Center for Global Health and Medicine; Takano M, Japanese Red Cross Musashino Hospital; Saito M, Yamada K, Takano H, The Jikei University School of Medicine; Toyoshima M, Nippon Medical School Hospital; Akira S, Meirikai Tokyo Yamato Hospital; Okada Y, St. Luke’s International Hospital; Nagai K, Miyagi E, Yokohama City University Hospital; Saito S, Yokohama City University Medical Center; Kuroda H, Kawasaki Saiwai Hospital; Sakamoto I, Yamanashi Prefectural Central Hospital; Horisawa S, Japanese Red Cross Society Nagano Hospital; Takahashi N, Shizuoka Cancer Center; Kosaka K, Shizuoka General Hospital; Itoh T, Hamamatsu University School of Medicine; Kobayashi M, Seirei Hamamatsu General Hospital; Otsuka K, Kasugai Municipal Hospital; Osafune A, Kariya Toyota General Hospital; Sawayama S, Sakurai A, Shiga General Hospital; Sunada M, Kyoto University Graduate School of Medicine; Matsumoto T, Osaka Central Hospital; Oki N, Chibune General Hospital; Yamabe E, Hyogo Prefectural Nishinomiya Hospital; Nagamata S, Terai Y, Kobe University Hospital; and Sato S, Oita Saiseikai Hita Hospital.

## Contribution to authorship

RK, ST, HT, and KM conceptualised and designed the study. RK, YHa, YHi, MN, ST, KS, EY, and HK collected and curated the data. RK, ST, and HM performed statistical analyses. ST secured funding for the study. RK, YHa, YHi, MN, ST, and HM carried out the investigation. RK, YS, and CK developed the methodology. ST supervised the project and led project administration. ST and HK were responsible for software implementation. RK, KS, EY, and HK provided essential resources. RK, ST, YS, and CK were involved in data visualisation. RK, ST, CK, HM, and KM drafted the manuscript. All authors critically reviewed and edited the manuscript; have read and approved the final version of the manuscript; meet the ICMJE criteria for authorship; and agree to be accountable for all aspects of the work.

## Details of ethics approval

This study was conducted in accordance with the principles embodied in the 1964 Declaration of Helsinki (as revised in Brazil in 2013). This study protocol was reviewed and approved by the Medical Research Ethics Review Committee of the Japanese Foundation for Cancer Research (registration no.: 2020-375) on 28 December 2020. Written informed consent was obtained from all patients for the use of their surgical videos and clinical data, including commercial use.

## Data availability

The data that support the findings of this study are available from the corresponding author upon reasonable request.

## Preprint notice

This manuscript has been submitted to BJOG and is currently under peer review. This preprint has not been certified by peer review.

